# Demographic science aids in understanding the spread and fatality rates of COVID-19

**DOI:** 10.1101/2020.03.15.20036293

**Authors:** Jennifer Beam Dowd, Liliana Andriano, Valentina Rotondi, David M. Brazel, Per Block, Xuejie Ding, Yan Liu, Melinda C. Mills

## Abstract

Governments around the world must rapidly mobilize and make difficult policy decisions to mitigate the COVID-19 pandemic. Because deaths have been concentrated at older ages, we highlight the important role of demography, particularly how the age structure of a population may help explain differences in fatality rates across countries and how transmission unfolds. We examine the role of age structure in deaths thus far in Italy and South Korea and illustrate how the pandemic could unfold in populations with similar population sizes but different age structures, showing a dramatically higher burden of mortality in countries with older versus younger populations. This powerful interaction of demography and current age-specific mortality for COVID-19 suggests that social distancing and other policies to slow transmission should consider both the age composition of local and national contexts as well as the social connectedness of older and younger generations. We also call for countries to provide case and fatality data disaggregated by age and sex to improve real-time targeted nowcasting.

## Background

Governments are rapidly mobilizing to minimize transmission of COVID-19 through social distancing and travel restrictions to reduce fatalities and the outstripping of healthcare capacity. It is becoming clear that the pandemic’s progression and impact may be strongly related to the demographic composition of the population, specifically population age structure. Demographic science can provide new insights into how the pandemic may unfold and the intensity and type of measures needed to slow it down.

Currently, COVID-19 mortality risk is highly concentrated at older ages, particularly those aged 80+. In China, case-fatality rate (CFR) estimates range from 0.4% (40-49 years), jumping to 14.8% (80+ years).(1) This is consistent with the data from Italy as of March 16, where the reported CFR is 11.8% for those 70-79, 18.8% for 80-89, and 21.6% for those >90. Thus far in Italy, only 1.5% of deaths have occurred in those under aged 60.(2) As testing becomes more widespread, these case fatality rates may come down due to an increase in the true denominator. Nonetheless, in South Korea where testing was broader and the health system not overtaxed, the current CFR for those 80+ is still an alarming 9.26%. (7)

### The Importance of Age Structure

Population age structure may explain the remarkable variation in fatalities across countries and why countries such as Italy are especially vulnerable. The deluge of critical and fatal COVID-19 cases in Italy was unexpected given the health and wealth of the affected region. Italy is one of the oldest populations in the world with 23.3% its population over age 65, compared to 12% in China (3). Italy is also a country characterized by extensive intergenerational contacts which are supported by a high degree of residential proximity between adult children and their parents (4). Even when inter-generational families do not live together, daily contacts among non-co-resident parent-child pairs are frequent. Many Italians also often prefer to live close to their extended family and commute to work daily. According to the latest available data by the Italian National Institute of Statistics, this extensive commuting affect over half of the population in the northern regions.(5) These intergenerational interactions, co-residence, and commuting patterns may have accelerated the outbreak in Italy through social networks that increased the proximity of elderly to initial cases (see Supplementary Information).

Age structure, along with early detection and treatment, also likely explains the low numbers of fatalities in South Korea and Singapore compared to Italy. The Korean outbreak, while large, was concentrated amongst the young recruits of the Shincheonji religious group (6), with only 3.3% of cases falling into the very vulnerable >80 group.(7) Singapore is notable with zero deaths thus far, but have had only one confirmed case over 80 and only 10/200 cases above age 70.(8)

COVID-19 transmission chains that begin in younger populations may have a low number of severe cases and thus go longer undetected, (9, 10) with countries thereby slow to raise the alarm. The low case fatality rate in England thus far may reflect the relatively young age structure of populations impacted to date, including Greater London, which has a small fraction of residents over age 65 compared to more rural areas (11). COVID-19 was only detected in King County, Washington once it reached the Life Care Centre in Kirkland, where 19 out of 22 deaths occurred, despite estimates based on virus genetic sequences suggesting it circulated for several weeks prior (12). Once community transmission is established, countries that have a high level of intergenerational contacts and co-residence may see faster transmission to high-fatality age groups as seen in Italy.

In Figure 1, we use population pyramids to illustrate how population age structure **interacts** with high COVID-19 mortality rates at older ages to generate large differences across populations in the number of deaths, using existing assumptions about infection prevalence and age-specific mortality. The top panel considers two countries currently affected, Italy and South Korea. The larger number of expected fatalities for Italy is clearly visible in the right panel. In the bottom panel, we consider two countries yet untouched by the pandemic who have similar population sizes but very different age distributions. In Brazil, which has 2.0% of its population age 80+, the simulated scenario leads dramatically more deaths (1,025,341) compared to Nigeria (300,715), where the fraction over 80 is only 0.2%. Figure 2 uses an alternative visualization to depict the expected deaths by age groups in Italy, Brazil, Nigeria, UK and US, together with the proportion of the population in different age groups. Both figures demonstrate the stark implications of an older population age structure for higher fatalities and critical cases.

**Figure 1.**
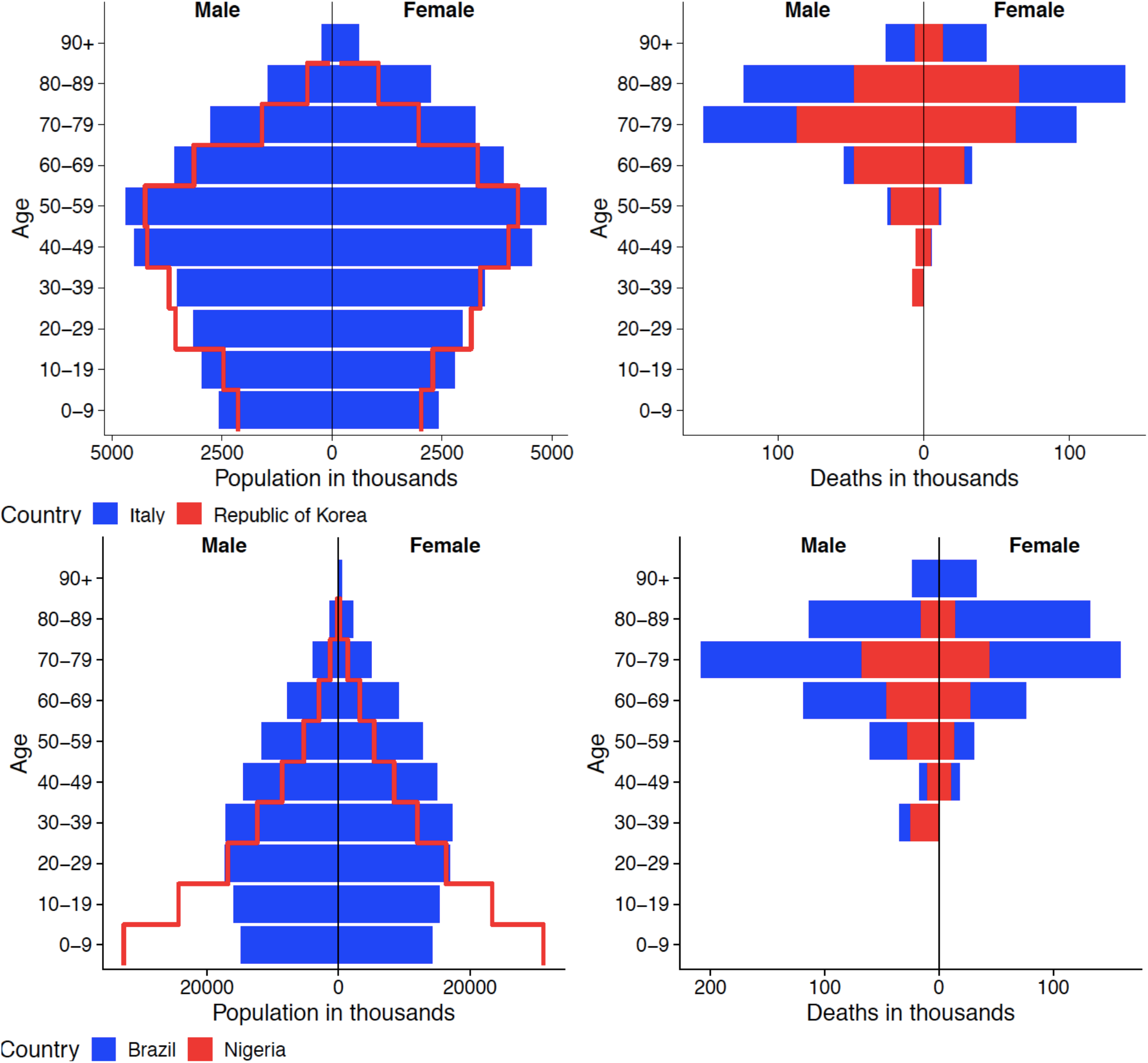
Population composition (left panel) and expected deaths in population (right panel), Italy and Republic of Korea (top panel) and Nigeria and Brazil (bottom panel)

**Figure 2.**
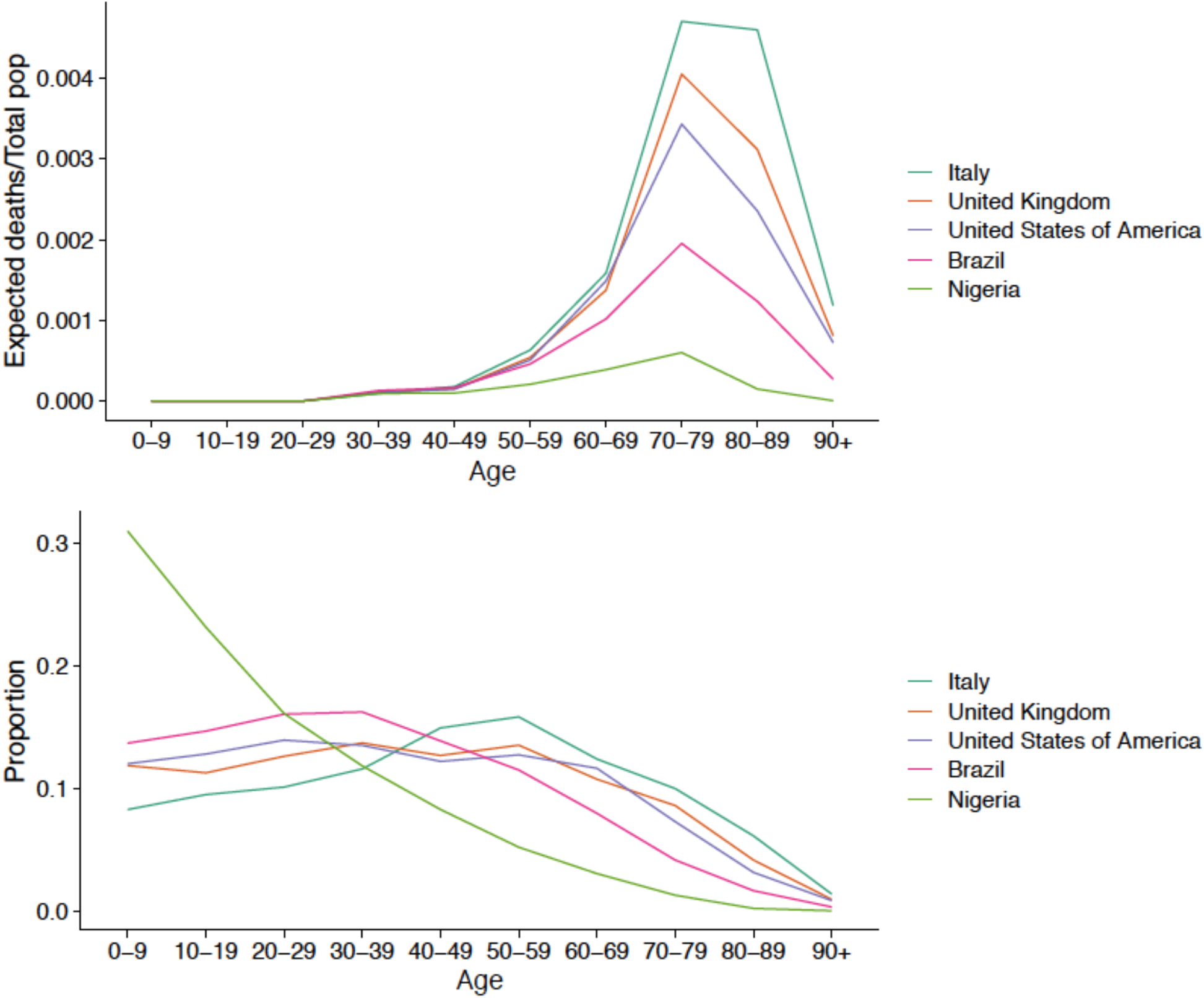
Expected deaths by total population (top panel) and proportion of total population by age group (bottom panel), Italy, United Kingdom, United States of America, Brazil and Nigeria

### Demographic Science and COVID-19 Policy

Going forward, demographically informed projections will better predict the COVID-19 burden and inform governments about targeted action. While population age structure is crucial for understanding the populations at the highest risk of mortality both across and within countries, it is also vital for understanding how much social distancing measures are required in each population to reduce the number of most critical cases and overload on the health system—aka “flattening the curve”(13). Our illustrations show that countries with older populations will need to take more aggressive protective measures to stay below the threshold of critical cases that outstrip health system capacity. For these measures to be effective, special attention should be devoted to those population groups that are more at risk and patterns of intergenerational contact.

Consideration of population age structure also necessitates understanding the interlinkage of policy measures and how one policy might create a domino effect of unintended consequences. While schools may be a hub of contact and virus transmission, school closings may inadvertently bring grandparents and children into closer contact if they become the default carers. In aged populations with close intergenerational ties, governments need to facilitate childcare solutions that reduce contact. In a pending decree, for instance, the Italian government will introduce a special leave for parents with children at home from school and a voucher (around 600 euros) for babysitting.

The age structure of populations also suggests that the often squeezed “sandwich” generation of adults who care for both the old and young are an important link for mitigating transmission. Beyond introducing sick pay for those who need to self-isolate or care for family members, joint government and industry emergency policy measures should seek to counter family economic crises by delaying rent and mortgage payments for example, particularly for vulnerable and precarious workers. In the absence of economic security measures, this crucial sandwich generation may be less able to comply with policies that allow social distancing.

## Conclusion

The rapid spread of COVID-19 has revealed the need to understand how population dynamics interact with pandemics now and in the future. Population ageing is currently more pronounced in wealthier countries, which mercifully may lessen the impact of this pandemic on poorer countries with weaker health systems but younger age structures. It is plausible that poor general health status and co-infections such as tuberculosis may still increase the danger of COVID-19 among younger cases in these countries. Thus far, the lower than expected number of cases detected in Africa (despite extensive trade and travel links with China), suggests that the young age structure of the continent may be protective of severe and thus detectable cases, or it may be undetected. Beyond age structure, there are large sex differences in mortality that need to be understood – with men at higher risk – some of which may be accounted for by the stark differences in smoking rates by sex in Asia. Distributions of underlying co-morbidities such as diabetes, hypertension and COPD will likewise refine risk estimates. Until these more nuanced data are available, the concentration of mortality risk in the oldest old ages remains one of the best tools we have to predict the burden of critical cases and thus more precise planning of availability of hospital beds, staff and other resources.

At this moment, few countries are routinely releasing their COVID-19 data with key demographic information such age, sex, or comorbidities. We call for the timely release of this disaggregated data to allow researchers and governments to nowcast risk for more focused prevention and preparedness.

## Methods

### Data

The data to produce Figures 1 and 2 use population data from (https://population.un.org/wpp) with the relative risk of death taken from the most recent Italian data, last accessed March 16, 2020, available in the Table S1.(14)

### Data Analysis

For Figures 1 and 2, the total number of expected deaths by age group was derived by multiplying the total number of people in each age-sex group and country by an assumed population infection rate of 0.4 and age-sex specific mortality rates extracted from most recent Italian data (March 16).

Data analysis is conducted in R using the packages [ggplot2].

## Data Availability

Population data is used from (https://population.un.org/wpp) with the relative risk of death taken from the most recent Italian data from Istituto Superiore di Sanità, last accessed March 13, 2020.(see ref 14) Supp Info analyses breakdown data across regions in Italy over time and geographically, with detailed data sources listed there.

## Author Contributions

All authors jointly devised the study. JBD & MCM drafted the manuscript in which all authors contributed and commented. VR & LA wrote the Italian case and graphics. PB worked on intergenerational transmission. LA and DMB generated graphics and with XD & YL, updated country statistics.

## Supplementary Information

### Demographic science aids in understanding the spread and fatality rates of COVID-19

#### 1. A Case Study of Population Ageing, Intergenerational Contacts, and COVID-19 in Italy

After Japan, Italy is currently the world’s second oldest population, with 23% of the population aged 65 years and older compared to 13.2% aged 15 and under.^1^ This population ageing and population decline has been driven by very low fertility rates and growing rates of childlessness,^1^ which are not compensated by immigration flows. In the ranking of current cases and deaths from coronavirus in the world, Italy sadly occupies the second position after China.

On Friday February 21^st^ 2020, the first case of COVID-19 in Italy was confirmed in in the province of Lodi, in Lombardy.^2^ Since then, other regions in Northern Italy – including Emilia Romagna, Piedmont, and Veneto – started to report rapid increases. Between February 24^th^ and March 13^th^, the number of cases and deaths in these regions increased exponentially (Figure S1).

The response of the Italian government to the spread of the coronavirus across Italy has been firm, yet not always effective. Most of the initial interventions targeted the four regions in northern Italy that were the most affected: Lombardy, Piedmont, Veneto, and Emilia Romagna. In particular, on February 23^rd^, the government issued a decree which prohibited the movement of people outside of 10 municipalities located in Lombardy, in the province of Lodi, and a municipality located in Veneto, in the province of Padova.^3^

While cases in the province of Bergamo began to increase from Feb 24th, in contrast to Lodi no shutdowns or restrictions were imposed. A few hours later, the Veneto and Lombardy regions issued an ordinance by which all schools of all levels and grades, including Universities, were closed and all cultural, recreational, sporting and religious events, being them public or private, were suspended.

Starting from March 8^th^, the shutdown implemented in the province of Lodi, was expanded to the entirety of Lombardy (including Bergamo) and to fourteen provinces in Veneto, Emilia Romagna, Piedmont, and Marche. The decree, however, leaked in the late evening of March 7^th^, generating a massive outflow of people from these regions in the North to several regions in the South, where the lowest number of cases are currently registered. Only two days later, a new decree of the Prime Minister, Giuseppe Conte, extended until April 3^rd^ identical measures to the entire country of Italy. On March 11^th^, a further crackdown resulted in the closure of all shops (except for groceries and pharmacies), pubs, and restaurants.

**Figure S1.**
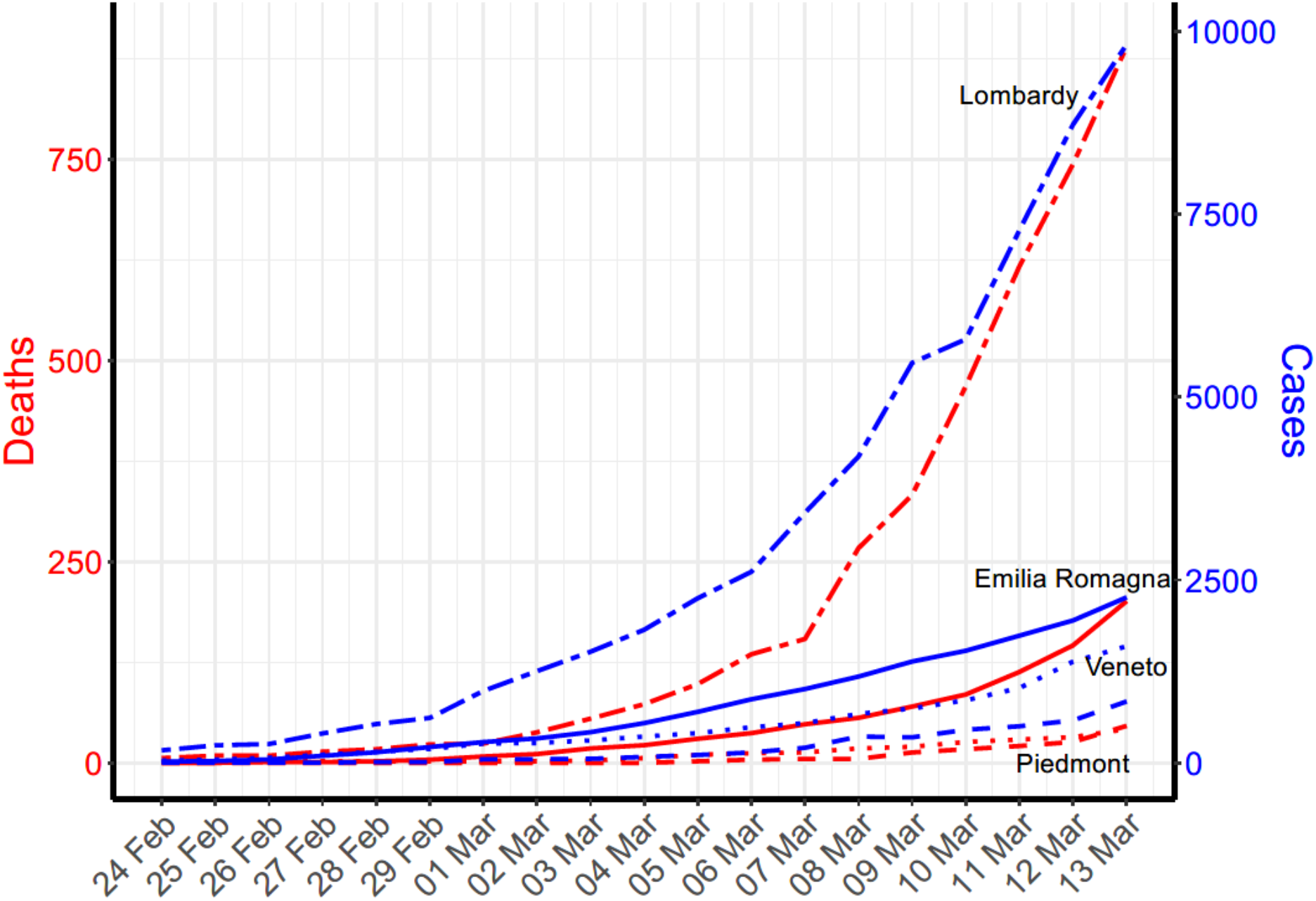
Number of cases and deaths across regions over time, March 13, 2020

As of March 13th, the number of cases that tested positive for COVID-19 in Italy amounted to 17,660, the number of deaths to 1,266 and the number of recovered to 1,439. With a total of 9,820 (i.e. 55.6% of the total) cases tested positive to COVID-19, 890 deaths, and 1,198 recovered, Lombardy is the most affected region in Italy (Figure S1).

Figure S2 illustrates the number of cases in Lombardy as of March 12^th^ by province. The more intense red indicates a greater number of cases, as reported in the legend.

**Figure S2.**
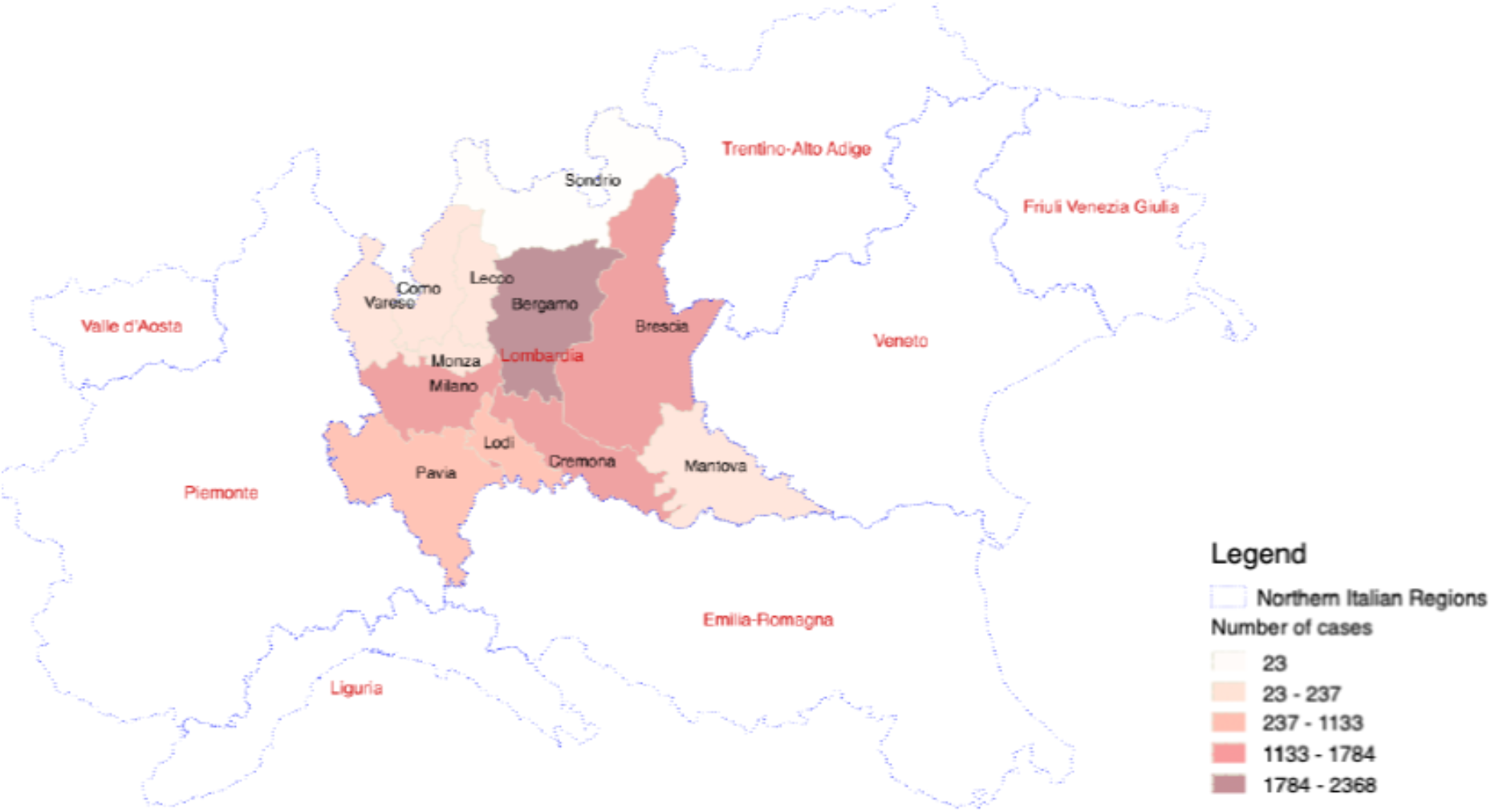
Number of cases by province as of March 13, 2020

As of March 13th, the most affected province of Bergamo (2,368 cases) has largely overcome the province of Lodi (1,133 cases) where the outbreak started and the containment measures were introduced first, as shown in Figure S3. We note that social distancing interventions were invoked on Feb 23^rd^ in Lodi but until March 8^th^ in Bergamo, providing some empirical evidence for the potential of “flattening the curve” interventions.

As already suggested, Italy is a country characterized by extensive intergenerational contacts which are supported by a high degree of residential proximity between adult children and their parents. Although proximity is the highest in smaller villages and in poorer regions, the geography of proximity across Italy is rather homogeneous, with the two richest regions — Lombardy and Trentino-Alto Adige — showing high rates of proximity as well. Even when inter-generational families do not live under the same roof, daily contacts among non-co-resident parent-child pairs are frequent. While on the one hand, this geographical proximity guarantees high rates of mutual intergenerational solidarity, both financial and in-kind, one obvious consequence is that Italians often prefer not to move for work but to live close to their family and commute daily to go to work. According to the latest available data by the Italian National Institute of Statistics, in the northern regions this extensive commuting affect over half of the population^4^.

**Figure S3.**
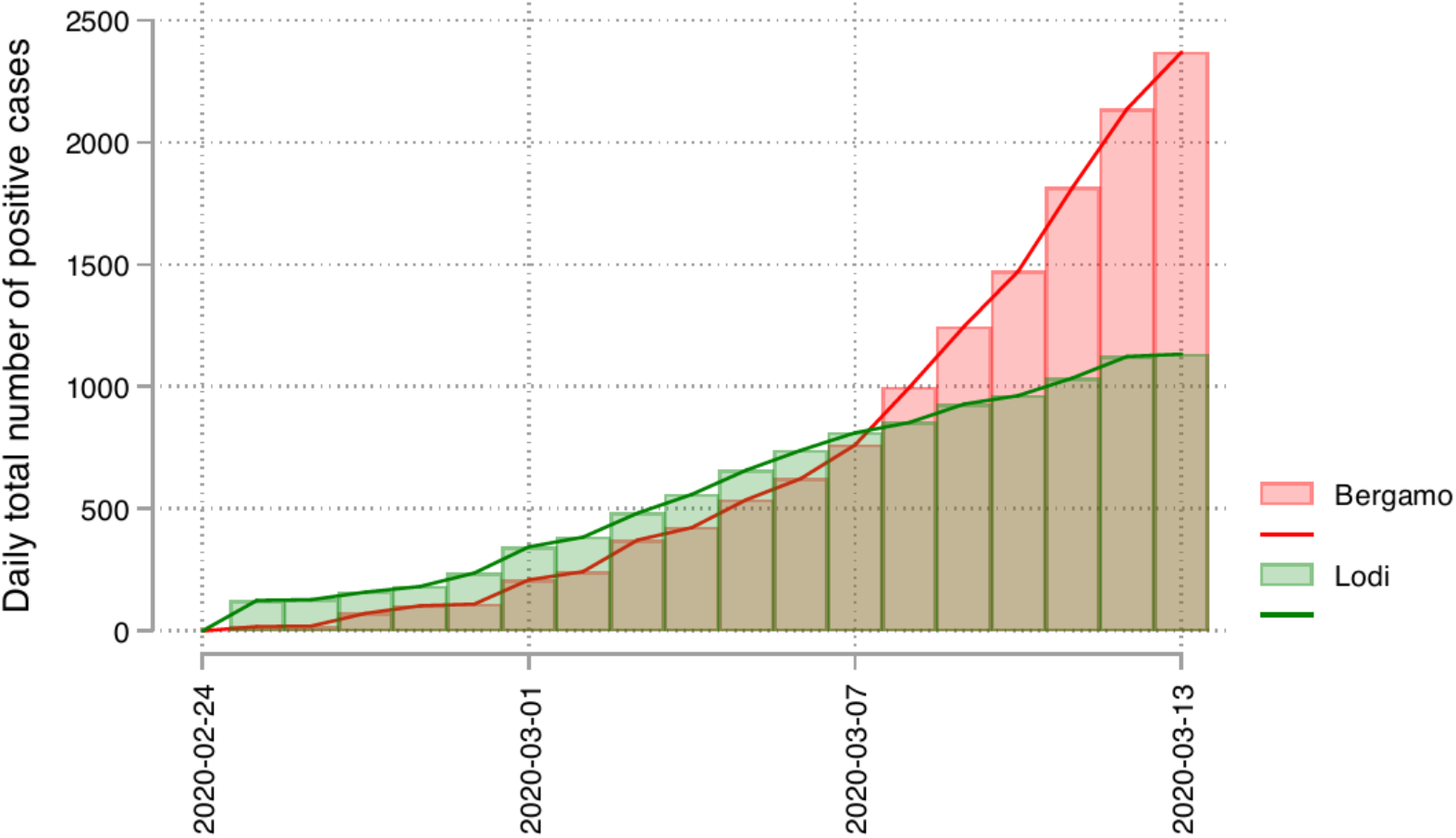
Number of cases in the Province of Bergamo (red) and Lodi (green) as of March 13, 2020

A stylized example from social network theory can be helpful in explaining why intergenerational interactions, co-residence, and commuting patterns might have played a role in the spread of the COVID-19 infection to the older population in Italy. Individuals’ social networks are generally composed of people similar in age. The population structure of contact can be represented as age-homogeneous communities that have low contact between groups. If the initial infections in northern Italy were younger people commuting to cities and having plausibly international contacts, a crucial determinant of risk for the elderly is their network distance to these younger sources, i.e. how many intermediaries need to be infected until they are reached. Network science showed that even relatively few connections between communities can lead to a stark reduction in average network distances; the so-called small world phenomenon^5^. Such community “connecting” individuals might be those young people around Milan that work in the city but reside in the most hard-hit villages in the surrounding with their parents and grandparents. Thus, intergenerational co-residence may have accelerated the outbreak by creating intercommunity connections that increase the proximity of elderly to the initial cases, an area for further study.

This stylized example may serve, once more, to show why while population age structure is crucial for understanding the populations at the highest risk of mortality both across and within countries, it is also important for understanding how much social distancing measures are required in each population to reduce the number of most critical cases and overload on the health system—aka “flattening the curve.”^6^ At this time of severe crisis, policy makers are called to define containment measures which are often difficult to sustain in the long run and which have immense repercussions in terms of socio-economic sustainability. For these measures to be effective, a special attention should be devoted to those population groups that are more at risk *and* to the strength of the connections across generations.

An interesting example in this direction comes from the Canton Ticino,^7^ the canton in Switzerland bordering to Lombardy. On March the 12^th^, the Canton Ticino has adopted measures aimed at contrasting the diffusion of the virus which are explicitly aimed at protecting the elderly and at-risk populations. To this aim, the resolution 1262^8^ “strongly discourages” people over 65 (and those categories at risk of incurring serious complications that can endanger their lives) to “look after children, participate in public or private events, use public transportation, except for medical and professional needs or for the purchase of basic necessities, and attend public exercise.” Further research to test how population age structure, intergenerational contacts, and social distancing measures interact to best mitigate risk is needed.

#### 2. Variation in fatality rates by different population structures

Here we replicate Figure 1 in Figures S4-S5 and show the variation in fatality rates with varying assumptions about the overall infection rate.

**Figure S4.**
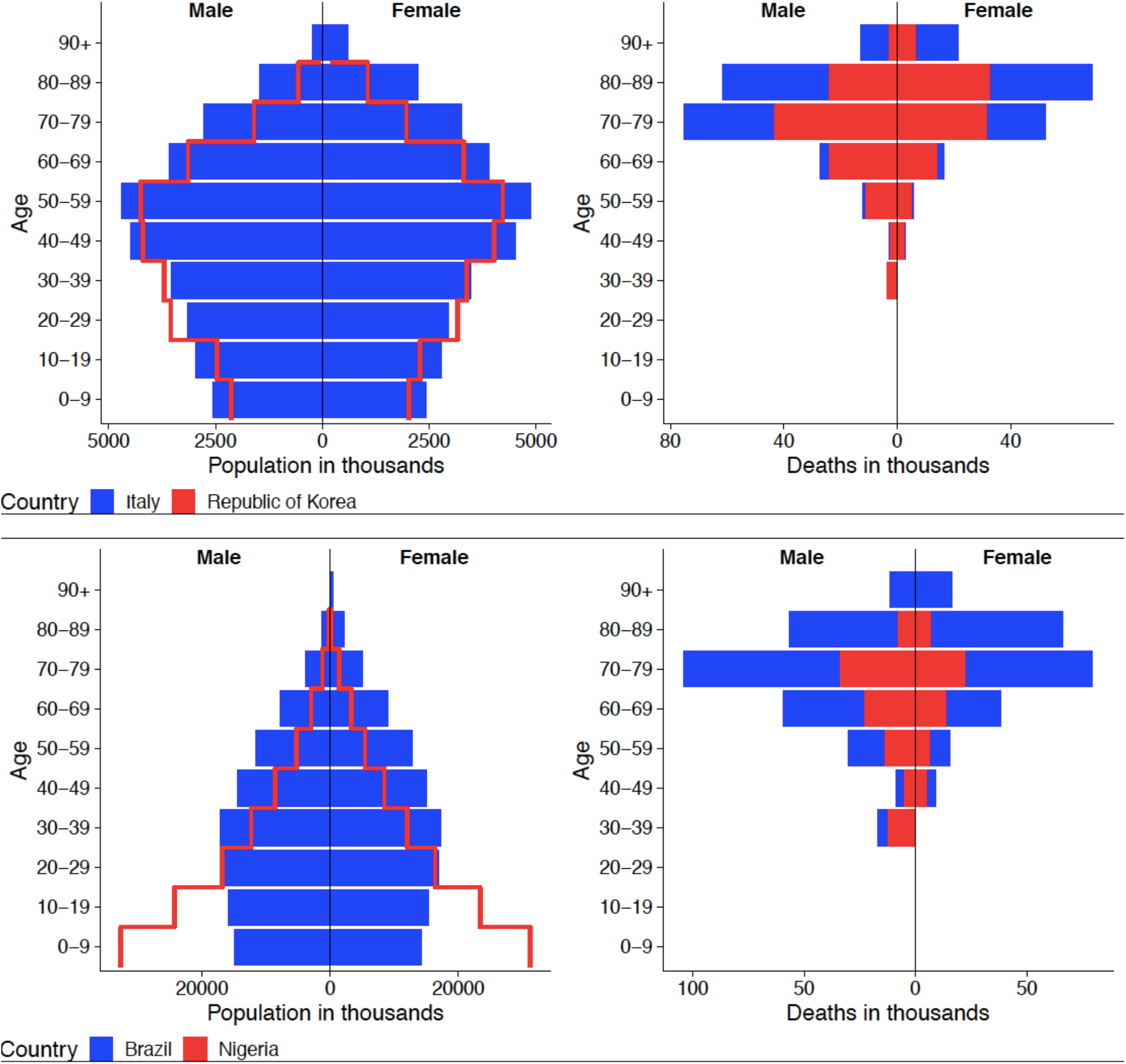
Population composition (left panel) and expected deaths in population (right panel), Italy and Republic of Korea (top panel) and Nigeria and Brazil (bottom panel) using Infection rate = 0.2; Note: Total number of expected deaths by age group is derived by multiplying the total number of people in each age group and country by an assumed infection rate of 0.2 and age-and sex-specific mortality rates extracted from most recent Italian data.

#### 3. Demographic population pyramid projections for additional countries

Figure S6 graphs the population composition and expected deaths in the population for the additional countries of the United States, Japan and South Africa. Japan has a relatively old population, South Africa a younger population and the US is more evenly distributed. Based on the age-specific mortality rates extracted from the Italian data, we project how these different countries will experience deaths attributed to COVID-19 by age and sex.

Figure S7 depicts the population composition (left panel) and the expected deaths in the population (right panel), this time for Italy versus the United Kingdom.

The United Kingdom is as of March 13 2020 is standing out as one of the few European countries to take not stringent actions such as closing schools or stopping large public events.^9^ In spite of the comparatively younger population of the UK, the bottom right panel illustrates that the UK could face similar numbers of COVID-19 deaths as Italy. Due to age structure differences, the UK will likely have slightly fewer deaths of those 80+ in comparison to Italy, but in the coming weeks still likely to face considerable pressure on its healthcare system.

**Figure S5.**
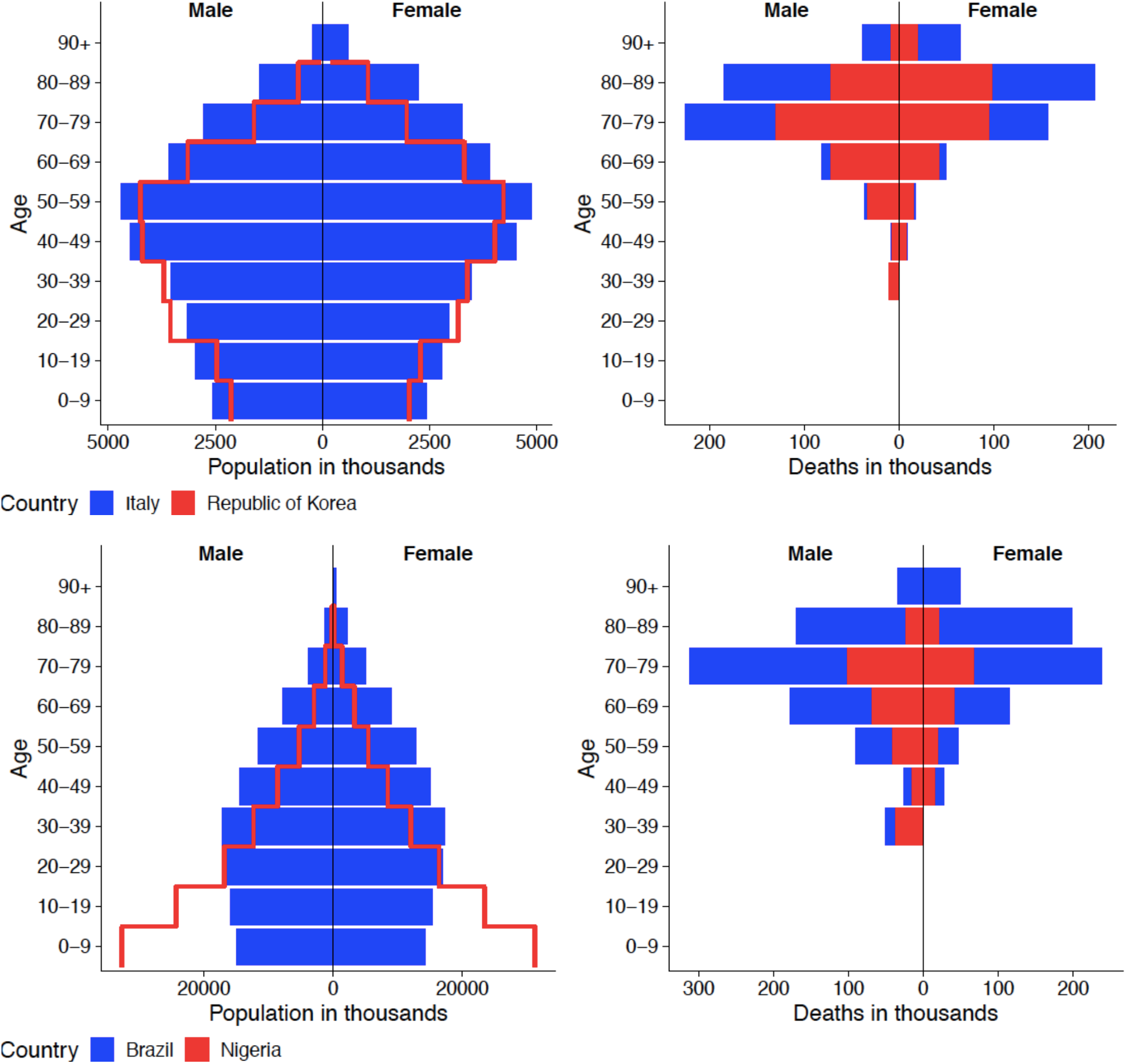
Population composition (left panel) and expected deaths in population (right panel), Italy and Republic of Korea (top panel) and Nigeria and Brazil (bottom panel) using Infection rate = 0.6; Note: Total number of expected deaths by age group is derived by multiplying the total number of people in each age group and country by an assumed infection rate of 0.2 and age-and sex-specific mortality rates extracted from most recent Italian data.

**Figure S6.**
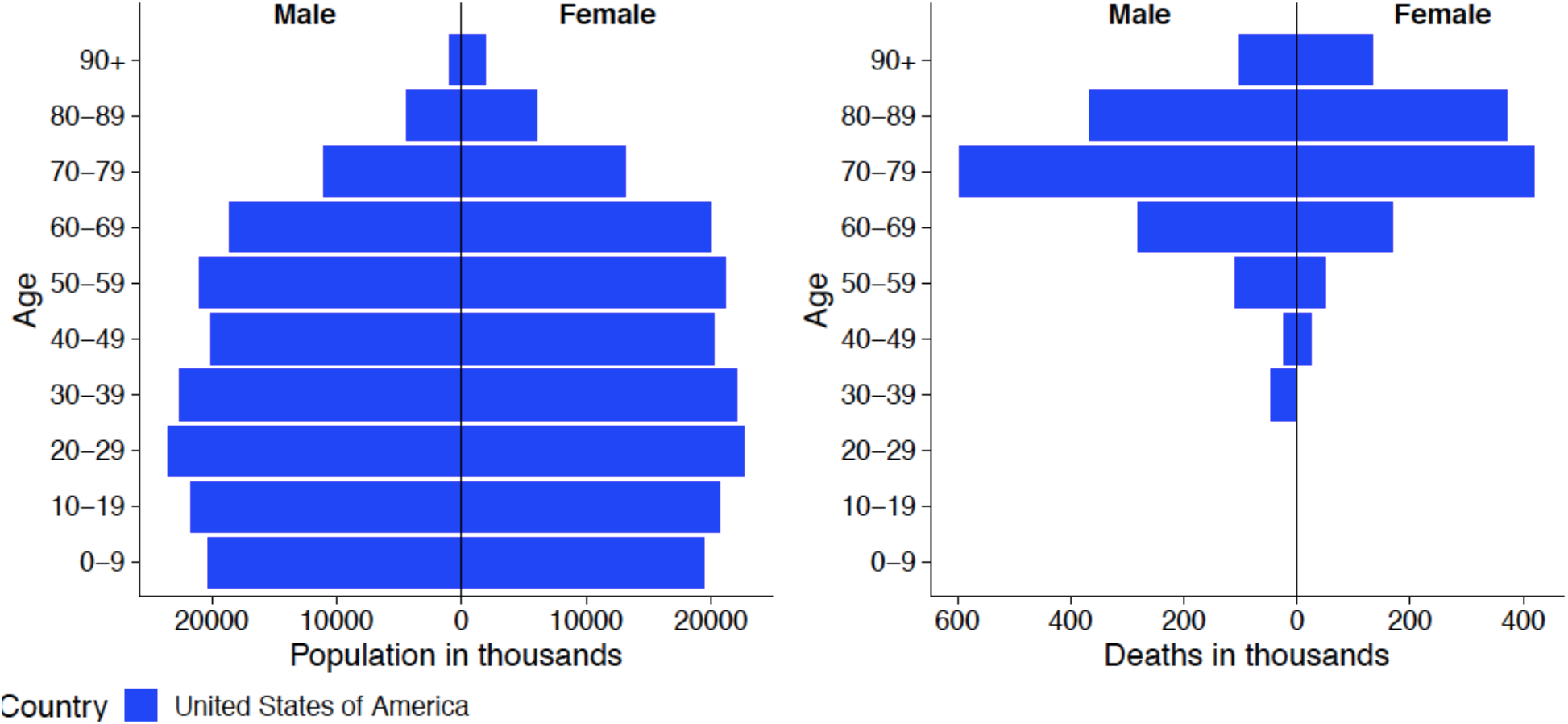
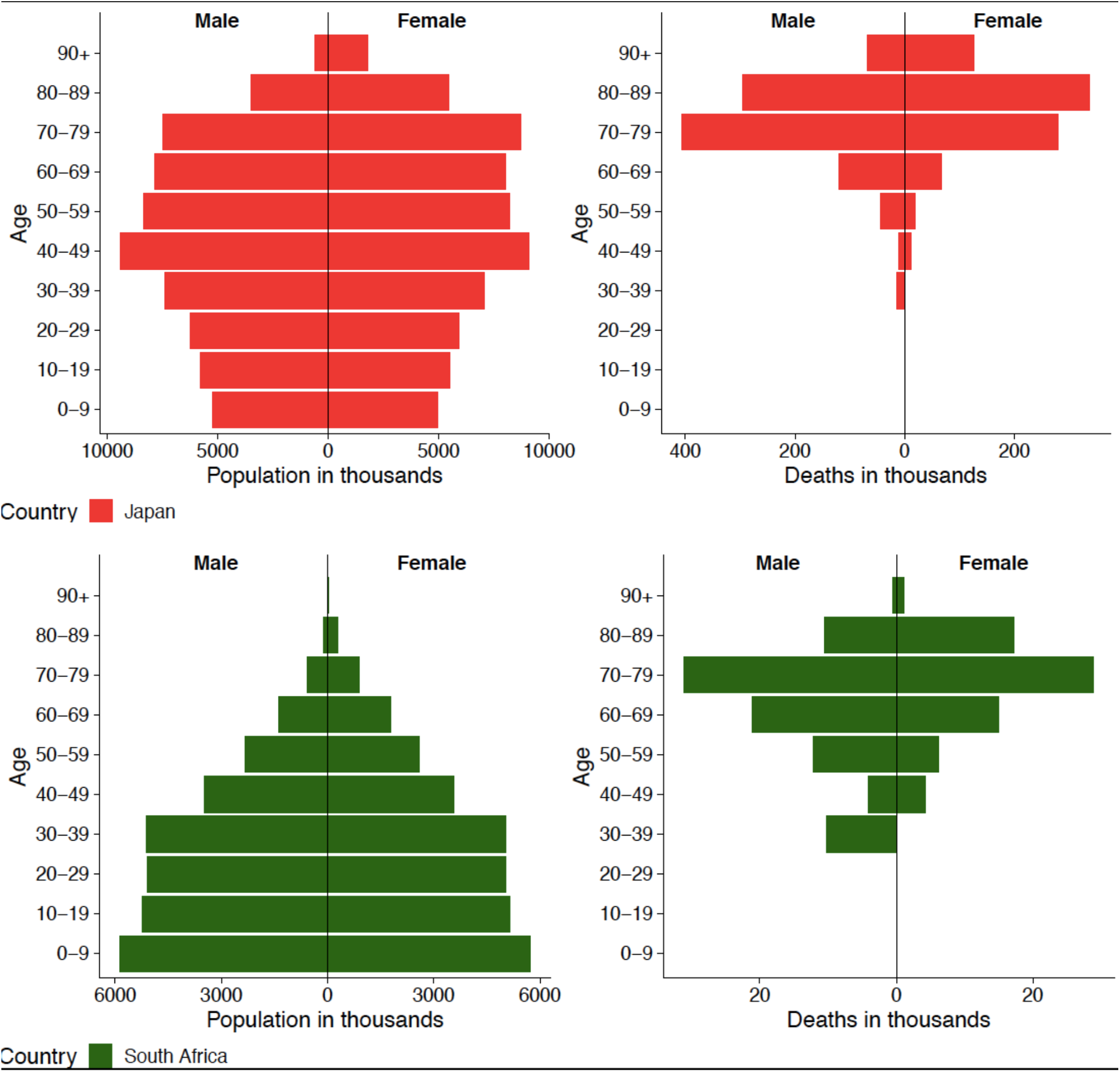
Population composition (left panel) and expected deaths in population (right panel), United States, Japan and South Africa Note: Total number of expected deaths by age group is derived by multiplying the total number of people in each age group and country by an assumed infection rate of 0.4 and age-and sex-specific mortality rates extracted from Italian data. The male-to-female relative risk of 1.65 based on current estimates from China.

**Figure S7.**
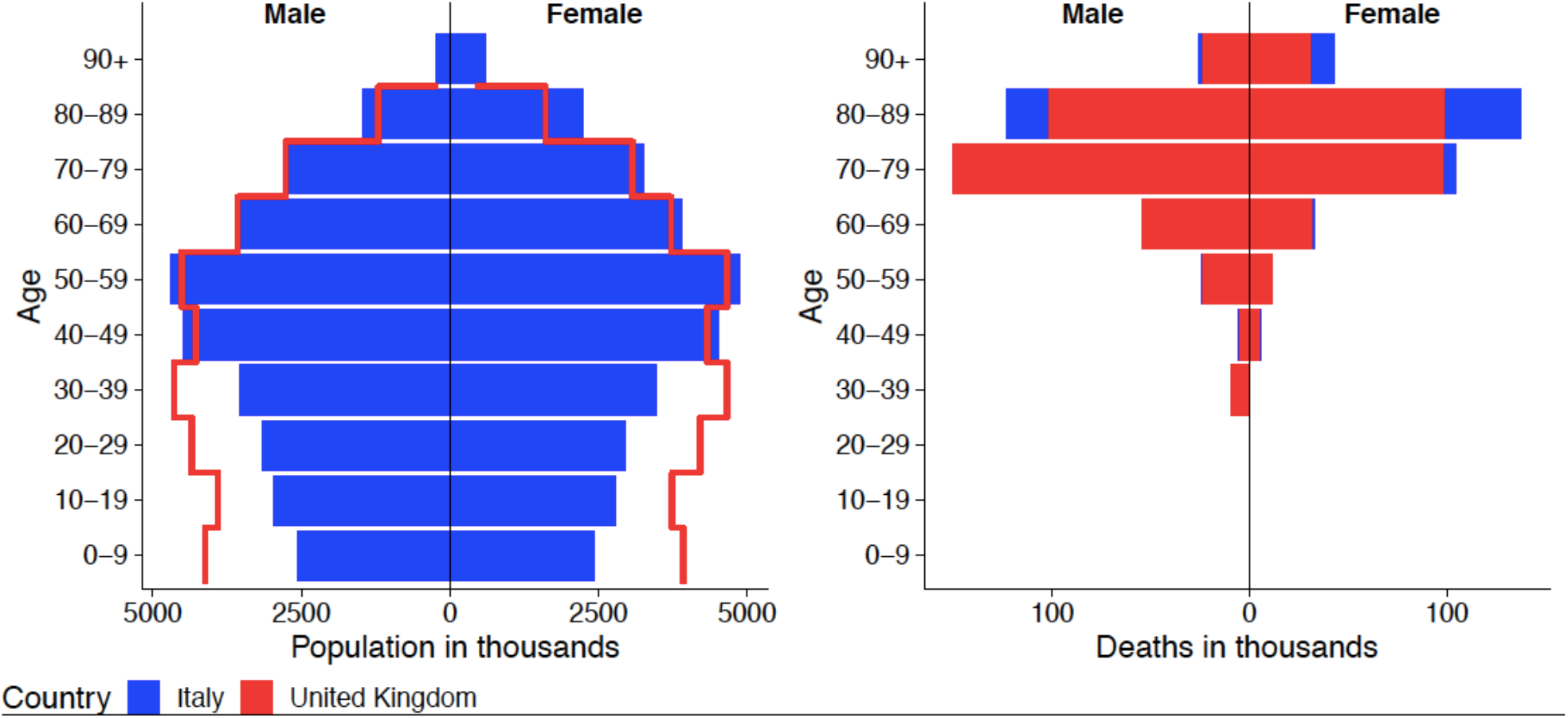
Population composition (left panel) and expected deaths in population (right panel), Italy and United Kingdom Note: Total number of expected deaths by age group is derived by multiplying the total number of people in each age group and country by an assumed infection rate of 0.4 and age-and sex-specific mortality rates extracted from Italian data.

**Table S1.** Age-Sex-Specific COVID-19 Case Fatality Rates from Italy as of March 16, 2020

| Table S1. Age-and-sex and case fatality rates, Italy (updated March 16, 2020) |  |  |  |  |  |  |  |  |  |  |  |  |  |  |  |
| --- | --- | --- | --- | --- | --- | --- | --- | --- | --- | --- | --- | --- | --- | --- | --- |
| Age group | Male |  |  |  |  | Female |  |  |  |  | Both |  |  |  |  |
|  | Number of | % of cases | Number of | % deaths | % fatality | Number of | % of cases | Number of | % deaths | % fatality | Number of | % of cases | Number of | % deaths | % fatality |
| 0-9 | 75 | 62.5 | 0 | 0 | 0 | 45 | 37.5 | 0 | 0 | 0 | 121 | 0.5 | 0 | 0 | 0 |
| 10-19 | 97 | 52.7 | 0 | 0 | 0 | 87 | 47.3 | 0 | 0 | 0 | 186 | 0.7 | 0 | 0 | 0 |
| 20-29 | 419 | 44.8 | 0 | 0 | 0 | 517 | 55.2 | 0 | 0 | 0 | 970 | 3.9 | 0 | 0 | 0 |
| 30-39 | 829 | 51.1 | 4 | 100 | 0.5 | 794 | 48.9 | 0 | 0 | 0 | 1,676 | 6.7 | 4 | 0.2 | 0.2 |
| 40-49 | 1,491 | 51.1 | 5 | 55.6 | 0.3 | 1,425 | 48.9 | 4 | 44.4 | 0.3 | 2,995 | 12.0 | 9 | 0.5 | 0.3 |
| 50-59 | 2,633 | 56.8 | 34 | 73.9 | 1.3 | 2,002 | 43.2 | 12 | 26.1 | 0.6 | 4,734 | 18.9 | 46 | 2.7 | 1.0 |
| 60-69 | 3,007 | 68.4 | 113 | 79.6 | 3.8 | 1,386 | 31.6 | 29 | 20.4 | 2.1 | 4,438 | 17.7 | 144 | 8.5 | 3.2 |
| 70-79 | 3,461 | 68.1 | 469 | 78.3 | 13.6 | 1,621 | 31.9 | 130 | 21.7 | 8.0 | 5,123 | 20.4 | 602 | 35.5 | 11.8 |
| 80-89 | 2,322 | 60.5 | 491 | 67.7 | 21.1 | 1,518 | 39.5 | 234 | 32.3 | 15.4 | 3,873 | 15.5 | 727 | 42.8 | 18.8 |
| 90+ | 287 | 37.8 | 81 | 49.1 | 28.2 | 472 | 62.2 | 84 | 50.9 | 17.8 | 763 | 3.0 | 165 | 9.7 | 21.6 |
| unknown | 99 | 57.2 | 0 | 0 | 0.0 | 74 | 42.8 | 0 | 0 | 0 | 179 | 0.7 | 0 | 0 | 0.0 |
| Total | 14,720 |  | 1,197 |  | 8.1 | 9,941 |  | 493 |  | 5.0 | 25,058 |  | 1,697 |  | 6.8 |

Source: Istituto Superiore di Sanità. Age specific mortality rates COVID-19 (2020) (March 16, 2020).
Source: Istituto Superiore di Sanità, Age specific mortality rates COVID-19 (2020) (March 16, 2020).
Source: Istituto Superiore di Sanità, Age specific mortality rates COVID-19 (2020) (March 16, 2020).

## Footnotes

1 Balbo N, Billari FC, Mills M. Fertility in Advanced Societies: A Review of Research. *Eur J Popul / Rev Eur Démographie* 2013; **29**: 1–38.

2 Lombardy is one of those Northern Italian regions that experienced a slight growth in the population in recent years. However, this growth is mainly due to immigration flows both from within and outside the country. Indeed, also in Lombardy the natural balance is negative and decreasing since 2012. According to Eurostat, Lombardy is also the most populated region in Italy and one among the wealthier regions in Europe, with a GDP per capita amounting to €37,800 in 2017 (compared to €28,400 in Italy and €29,500 in the EU) (https://ec.europa.eu/growth/tools-databases/regional-innovation-monitor/base-profile/lombardy)

3 However, the decree did not have immediate implementation. The control plan did not start rigidly, and checkpoints were not very effective. Law enforcement officers (at least 500 men) are deployed only on February 25 to ensure that no one enters and exits the so-called red areas creating 35 gates.

4 https://www.istat.it/it/archivio/224469

5 Watts, D. J. (1999). “Networks, dynamics, and the small-world phenomenon.” *American Journal of Sociology* **105**(2): 493-527.

6 https://www.nytimes.com/2020/03/11/science/coronavirus-curve-mitigation-infection.html

7 We thank Prof. Luca Crivelli for this suggestion.

8 https://www4.ti.ch/fileadmin/DSS/DSP/UMC/malattie_infettive/Coronavirus/1262.pdf

9 https://en.unesco.org/themes/education-emergencies/coronavirus-school-closures

